# *CovidNLP*: A Web Application for Distilling Systemic Implications of COVID-19 Pandemic with Natural Language Processing

**DOI:** 10.1101/2020.04.25.20079129

**Authors:** Raghav Awasthi, Ridam Pal, Pradeep Singh, Aditya Nagori, Suryatej Reddy, Amogh Gulati, Ponnurangam Kumaraguru, Tavpritesh Sethi

## Abstract

The flood of conflicting COVID-19 research has revealed that COVID-19 continues to be an enigma. Although more than 14,000 research articles on COVID-19 have been published with the disease taking a pandemic proportion, clinicians and researchers are struggling to distill knowledge for furthering clinical management and research. In this study, we address this gap for a targeted user group, i.e. clinicians, researchers, and policymakers by applying natural language processing to develop a *CovidNLP* dashboard in order to speed up knowledge discovery. The WHO has created a repository of about more than 5000 peer-reviewed and curated research articles on varied aspects including epidemiology, clinical features, diagnosis, treatment, social factors, and economics. We summarised all the articles in the WHO Database through an extractive summarizer followed by an exploration of the feature space using word embeddings which were then used to visualize the summarized associations of COVID-19 as found in the text. Clinicians, researchers, and policymakers will not only discover the direct effects of COVID-19 but also the systematic implications such as the anticipated rise in TB and cancer mortality due to the non-availability of drugs during the export lockdown as highlighted by our models. These demonstrate the utility of mining massive literature with natural language processing for rapid distillation and knowledge updates. This can help the users understand, synthesize, and take pre-emptive action with the available peer-reviewed evidence on COVID-19. Our models will be continuously updated with new literature and we have made our resource *CovidNLP* publicly available in a user-friendly fashion at http://covidnlp.tavlab.iiitd.edu.in/.

**Data Availability Statement:** All the data used in this study are publicly available from the WHO Covid-19 Global Literature on coronavirus disease maintained at https://search.bvsalud.org/global-literature-on-novel-coronavirus-2019-ncov/. Our analysis and the interactive resource *CovidNLP* is publicly available in a user friendly fashion at http://covidnlp.tavlab.iiitd.edu.in

## Introduction

Since 31st december 2019 when the first covid-19 case was reported from Wuhan city of China, more than 200 countries have been affected by the pandemic. Around 200,000 confirmed cases with more than 100,000 deaths have been recorded **[4]**. Clinical presentation is diverse, evidence for effective drugs is scanty, vaccine development can take much longer and systemic implications of the pandemic are unclear. However, more than 14,000 research articles peer reviewed/ preprint are publicly available **[5]** which can aid fast-tracking of knowledge synthesis to address these gaps. WHO alone has collated a list of more than 5000 articles so far which is expected to increase over the coming weeks. Synthesizing knowledge from peer-reviewed literature is not easy for the consumers of this information, i.e. clinicians, researchers and policymakers.

Furthermore, the re-allocation of resources towards COVID-19 is not without systemic consequences. It can be anticipated that in the near future, the mortality and rate of complications arising from other diseases, treatment for which has been put on hold, may become as significant as the COVID-19 itself. Hence it is important to have an updated system for knowledge synthesis and evidence to guide clinical policy. Delay in detection and treatment of many conditions such as cancer and tuberculosis may have long term health and economic consequences that will become evident as the acute crisis settles down. Since it is humanly not possible to consider all aspects, evidence based insights and knowledge synthesis will be critical to guide the health systems towards recovery. However we did not find any study that evaluates the emerging systemic effects and the extent of collateral damage that COVID-19 incurs. Hence we carried out a study exploring the role of word-embedding models, text summarization and interactive dashboards in guiding policy.

Artificial Intelligence and Natural Language Processing can provide novel insights to aid the distillation of insights from these data as healthcare personnel, researchers and policymakers are getting overwhelmed with data. We address this gap by using natural language processing and machine learning upon peer-reviewed research articles curated by the WHO, in order to discover hidden knowledge that can guide COVID-19 policy, research and development. We summarized these articles using text-summarization approaches and the trained word-embedding models. Exploring the space of words which has been used in the abstract of articles and identified the spectrum of clinical and non-clinical associations of COVID-19. Finally, we present this analysis as *CovidNLP*, an open-source dashboard that will be updated with new insights as the pre-trained models ingest more literature across different dimensions of health, social factors, economics and policy.

## Methods

### Data sets

Literature from the **WHO Database[6]** which contains more than 5363 peer-reviewed and published research articles till 13th april 2020 was downloaded.

### Pre-processing

Standard pre-processing steps such as lowercase conversion and removal of white-spaces, punctuation, stop words, digits and date-time information were carried out.

### Exploratory data analysis

A word cloud visualization of abstracts was carried out. Word clouds are a graphical representation of the word-frequency in a text and provide a first impression of the text documents in the corpus [7]. The font size, orientation, position, colour, repetition etc are used to encode basic statistics for the corpus information[8]. EDA was also carried out to understand the patterns of publication and venues.

### Word-embedding models

A low-dimensional representation of the corpus trained through word2Vec algorithm[34] which was then visualized in 3 dimensions in order to aid the exploration of feature space. word2Vec is a two layered neural network, where the neurons are trained over the corpus to capture the context of each word. Words with similar context tend to have similar embeddings vectors thereby capturing the relationship between them while iterating through large corpus. Skip-gram algorithm[45] was used and the neural network was trained with one-hot encoded vectors to predict the target word with a fixed window size, iterating along the corpus. Softmax function in the last layer computed the probabilities, and the errors against actual class labels were used to update weights in the backpropagation. The weight matrix hence computed generates word embeddings when multiplied with the one hot-vectors of words.

### Text summarization

Latent Semantic Analysis (LSA), an unsupervised learning method, was performed for extractive summarization of abstracts [9,10]. Briefly, an n × m term-sentence matrix (n terms, m sentences) was constructed with weights populated from using the Term Frequency-Inverse Document Frequency method. Singular Value Decomposition (SVD) was used to decompose the matrix into a term topic matrix, and a topic sentence matrix. The diagonal matrix encodes the weight of topics along the diagonal and defines how much a sentence resembles a topic.

### Model Inference

The space of lower dimensional representation was explored using Tensorflow Projector and was used for interactive construction of word-algebra queries. Only the top 100 words were preserved for querying the model and to construct inferences.

### *CovidNLP* Dashboard

Users can interactively query our models and visualize insights to form hypotheses on *CovidNLP* which is publicly available at http://covidnlp.tavlab.iiitd.edu.in/. These models will be continuously updated with new peer-reviewed literature as it becomes published.

## Results

The main motive of this study was to inform and query the systemic implications of COVID-19 on a dashboard that ingests peer-reviewed literature, summarizes findings, learns word-embedding models and allows interactive explorations on a daily basis. The dashboard, *CovidNLP* is made openly available to the public.

### Exploratory Data Analysis

The acceptance patterns of COVID-19 research by journals. We calculated the count of papers published in different journals, we found BMJ has accepted the highest research article with count 173.

### Word Embedding and cloud of words

Preliminary impressions from word cloud analysis revealed SARS CoV and COVID as the most frequently used words in the peer reviewed literature. These are presented to the user to aid effective queries. Word cloud analysis was used to get preliminary impressions about the co-occurrences of COVID related words of which “n-covid” was the highest frequency [fig2]. We then used embedding generated by word2vec to explore the space of words and by using cosine similarity identified the disease related words which came most similar with the word “n-covid”[Table 2].

**Table 1.** Words most similar to the word “n-covid” with cosine similarity. These keywords were then used to funnel the articles for text summarization in the pipeline.

| Word | Similarity | Word | Similarity | Word | Similarity | Word | Similarity |
| --- | --- | --- | --- | --- | --- | --- | --- |
| Renal | 0.9448740482 | palpitation | 0.9015104771 | refractory | 0.8795773387 | malignancy | 0.8494654298 |
| Fevers | 0.9439075589 | radiographic | 0.8999356627 | miscarriage | 0.8759618998 | tomography | 0.8454025984 |
| Diarrhoea | 0.9129174352 | nausea | 0.8964273334 | cardiac | 0.8756124377 | hypotension | 0.8418673277 |
| Comorbidity | 0.9023435116 | pneumonias | 0.8904905915 | intravascular | 0.8735768199 | encephalitis | 0.8330945373 |

**Table 2:** Count of articles having keywords like disease/risk in their articles identified using the word-embeddings. CovidNLP funnels the identified research into the text-summarization pipeline for easy knowledge distillation by the users (researchers, clinicians and policymakers)

| Category | Key words | Article Count |
| --- | --- | --- |
| Respiratory Disease | Pneumonia<br>CardioVascular<br>Rheumatic | 403<br>56<br>5 |
| Infectious Disease | HIV<br>Diarrhea<br>Tuberculosis | 19<br>6<br>6 |
| Maternal & Child Health | Pregnant | 35 |
| Cancer | Cancer | 105 |
| Risk | Risk | 137 |

**Fig 1:**
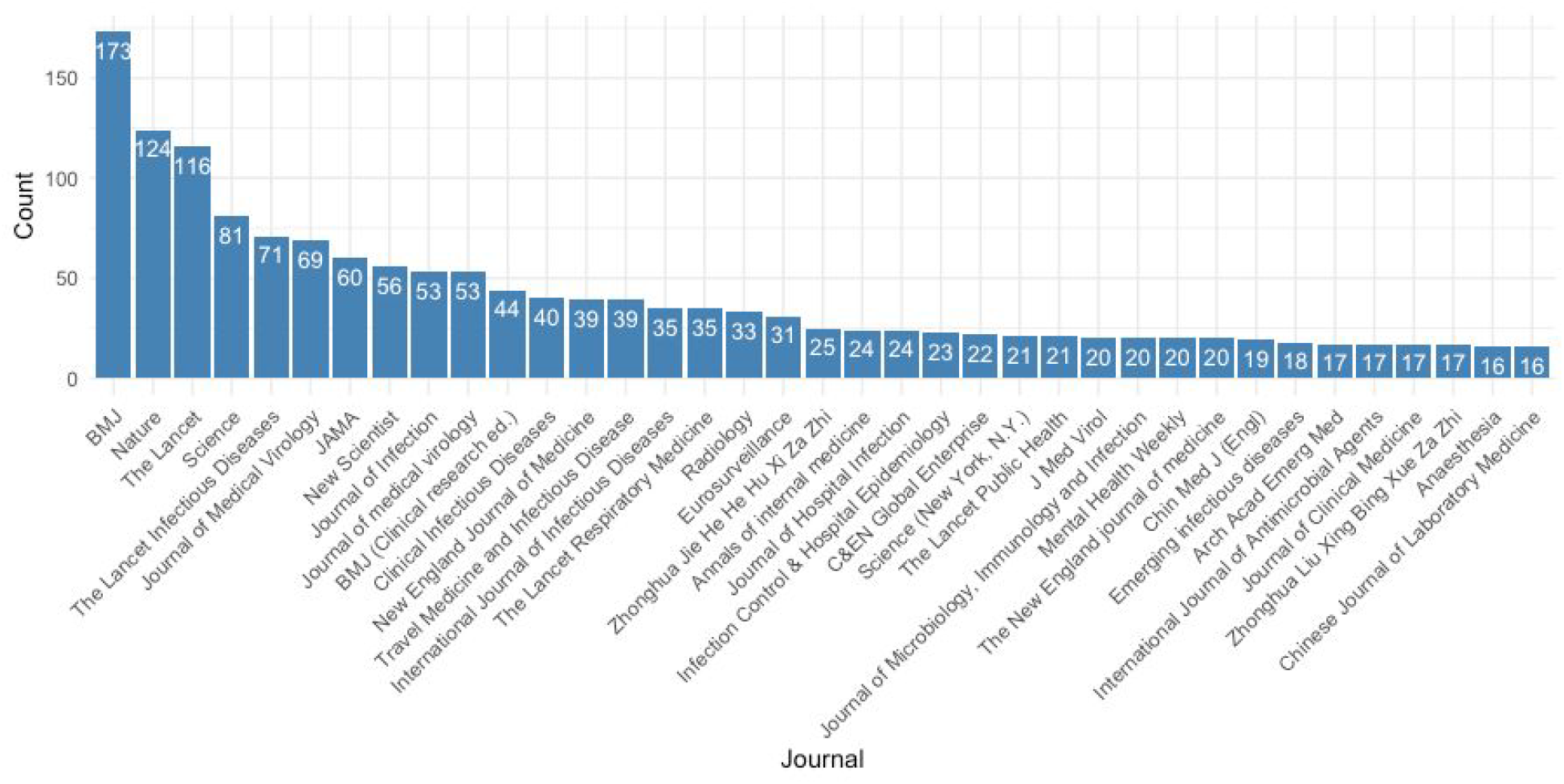
Bar plot shows count of published papers in different journals here we plotted only journals which have more than 15 papers.

**Fig 2:**
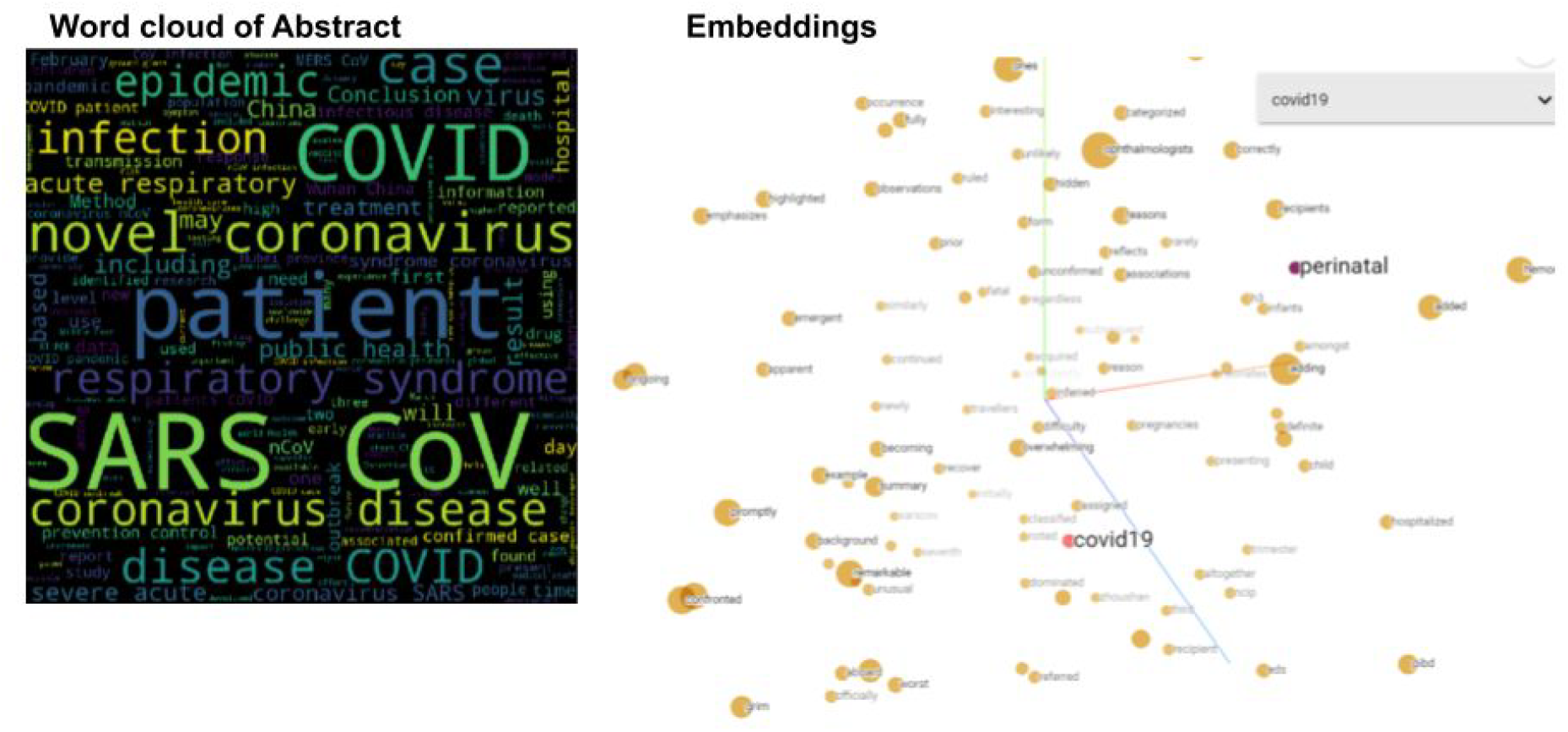
**Word** cloud generated by frequency count of words in the abstract of title and embedding of words generated by word2Vec and visualized on the CovidNLP dashboard.

### User Workflow and a Case Study on COVID Spectrum using *CovidNLP*

#### Word Embeddings based reconstruction of COVID-19 Spectrum

Using the word space and expert knowledge, a user can interactively query the *CovidNLP* dashboard for insights. An example use-case for exploration of the disease spectrum COVID-19 was created[table2]. These keywords were then used to guide the identification of articles that were then funneled into the text-summarization pipeline.

#### Extractive Summarization of Information about the Disease Spectrum

The user may get a summarized content filtered by the interactive querying of the dashboard. Our text summarization approach revealed interesting features of COVID which may be used to provide summary information from latest research to frontline health workers. Some of the key contextual features highlighted by the model are as follows:-

1. **Pneumonia**:
  - A collection of 403 papers associated with pneumonia in the database; we summarise below the abstract of these articles into 10 sentences and listed meaningful points:
  - In comparison to normal patients COVID-19 patients had lower counts of white blood cells, neutrophils, C-reactive protein (CRP), and alanine aminotransferase (ALT) on admission[17].
  - Interestingly, after lopinavir/ritonavir (Kaletra, AbbVie) was administered, ß-coronavirus viral loads significantly decreased and no or little coronavirus titers were observed.Background Chest CT is used to assess the severity of lung involvement in COVID-19 pneumonia[38].
  - Novel coronavirus pneumonia combined with liver damage is more likely to be caused by adverse drug reactions and systemic inflammation in severe patients receiving medical treatment[39].
  - We report a case where serial follow-up chest computed tomography revealed progression of pulmonary lesions into confluent bilateral consolidation with lower lung predominance, thereby confirming COVID-19 pneumonia[40].
  - We aim to define the chest computed tomography findings of 2019-novel coronavirus associated with pneumonia and its successful resolution after treatment[41].
  - The results of this multicenter retrospective study suggests that novel coronavirus pneumonia combined with liver damage is more likely to be caused by adverse drug reactions and systemic inflammation in severe patients receiving medical treatment[39].
2. **Tuberculosis**: A Collection of six papers got associated with the disease COVID in the vocabulary of titles thereby stating about its low significance/association with the disease. Key factors that got highlighted states how the export of the drugs for tuberculosis got hindered amidly due to the lockdown and ceased flight movements as stated by U.S. based humanitarian organization officials. Officials also suggested that North Korea is going to run out of TB-drug by May or June, which will be extremely hazardous for patients infected with multidrug-resistant TB strains[11].
3. **HIV:** A collection of 19 papers were extracted where both the diseases HIV and COVID have shown its co-occurrence in the title. Insights depicted about decrement in HIV testing which is the stepping stone for HIV care. This is affected due to lockdown, quarantine and social distancing, hampering UNAIDS’ first 90-90-90 target at a global sphere which states how 90% HIV-positive patients will be verified about their HIV status[36].
4. **Cancer:**
  - There were 105 papers associated with cancers in the database. We summarise the abstract of these articles into 15 sentences and listed meaningful key points below:
  - Results In a risk-mitigation pandemic scenario, efforts should be made not to compromise the prognosis of lung cancer patients by departing from guideline-recommended radiotherapy practice[12].
  - A few reports have shown that mortality due to SARS-CoV-2 is higher in elderly patients and other active comorbidities including cancer[13].
  - Oncologists should be more attentive to detect coronavirus infection early, as any type of advanced cancer is at much higher risk for unfavorable outcomes[14].
  - Given the systemic immunosuppressive state caused by malignancy and anticancer treatments, patients with advanced lung cancer may be at a higher risk of COVID-19 infection[15].
  - Results of MTT assay revealed that seven synthesized compounds (i.e., 11a, 11b, 12a, 12b, 13b, 13c and 13h) particularly exhibited significant cytotoxicity against the three cancerous cell lines under investigation[16].
5. **Maternal & Child Health**: There were 35 papers associated with the word Pregnant in the database. The summarized abstracts and meaningful key points are listed below:
  - No evidence of complications during veginal and C-section delivery by covid-19 infected pregnant women was observed[17].
  - From covid-19 pregnant women no evidence was found for intrauterine and trans placental transmission to their fetuses[18][20].

#### *CovidNLP* pipeline for risk identification

To understand the risk from the literature we filter the article using the keyword ***risk*** and we found 137 titles talking about the risk we summarise the abstract of all filtered articles into 15 sentences. followings are meaningful points with the summary

- Health promotion strategies for students.,COVID-19 is a devastating global pandemic with epicenters in China, Italy, Spain, and now the United States[21].
- Published findings indicate that SARS-CoV can enter the human nervous system with evidence from both postmortem brains and detection in cerebrospinal fluid of infected individuals[21].
- Endoscopists face risk for infection with viruses like SARS-CoV-2, as the aerosol generating nature of endoscopy diffuses respiratory disease that can be spread via an airborne and droplet route[22].
- This intriguing finding emerged from several studies that examined underlying medical conditions in COVID-19 patients.,The COVID-19 pandemic demands reassessment of head and neck oncology treatment paradigms[23].

#### *CovidNLP* pipeline for treatment implications

Clinicians and hospital policymakers can synthesize evidence for better management of COVID using the outlined methods and dashboard. The common questions addressed belong to diagnosis, prognosis and treatment strategies.

#### Diagnosis & Prognosis

We found 108 papers with the keyword “Diagnosis” and 15 papers with “prognosis” in their title, summary of abstract of these papers :

- The most reported predictors of severe prognosis in patients with covid-19 included age, sex, features derived from computed tomography scans, C reactive protein, lactic dehydrogenase, and lymphocyte count[24].
- We hope our “Renji experience” will be beneficial to colleagues.,Since December 2019, China has been experiencing an outbreak of new infectious disease caused by the 2019 novel coronavirus (2019-nCoV)[25].
- retrospective study in 12 cases with COVID-19 who underwent endotracheal intubation at ICU of the Guangzhou eighth hospital from January 20 to February 10, 2020[26].
- It is particularly important for medical institutions of all levels to maintain safe and effective routine services while doing well in COVID-19 prevention[27].
- Immediate sharing of well documented individual participant data from covid-19 studies is needed for collaborative efforts to develop more rigorous prediction models and validate existing ones[28].
- Methodological guidance should be followed because unreliable predictions could cause more harm than benefit in guiding clinical decisions[28].

#### The Hydroxychloroquine controversy

Hydroxychloroquine has been suggested as a potential treatment but evidence to date does not offer clarity on its efficacy. On exploration, 29 articles were found having Hydroxychloroquine as a keyword in their article, which are summarized as follows:

- Two antimalarial agents, chloroquine (CQ) and hydroxychloroquine (HCQ), have been trusted treatments for a range of rheumatic diseases over the past seventy years[42]
- all ongoing clinical trials with HCQ use different dosing regimens, resulting on various concentrations PK studies are therefore needed to define the optimal dosing regimen.A severe cutaneous drug reaction resembling acute generalized exanthematous pustulosis resulting from ingestion of hydroxychloroquine has been documented[43].

#### Efficacy of Remdesivir

Remdesivir is another drug which has been suggested as a treatment. Fifteen articles were found having Remdesivir as a keyword in their article summary of abstract of these papers:

- Remdesivir (RDV) is an investigational compound with broad spectrum of antiviral activities against RNA viruses, including SARS-CoV and Middle East respiratory syndrome (MERS-CoV) [44].
- The selectivity value for RDV-TP obtained here with a steady-state approach suggests that it is more efficiently incorporated than ATP and two other nucleotide analogues [44].

#### Systemic implications: Social, Economic and Education

Other than disease and treatment level terms we filtered the paper title using the words like “Education”, “Economy”, “Finance”, “Social Distancing”, and “Quarantine” which may indicate systemic consequences of COVID-19 and factors of relevance to the policymaker.

### Economy

Four papers were found having Economy keyword in their title, summary of abstract of these papers :

- It concludes by stating why preparedness for and response to all disease outbreaks, also in countries of lower economic importance, should become a priority in the global health agenda.[29].
- It then uses the trend comparison method to predict the inflection point and Key Point of the COVID-19 virus by comparison with the severe acute respiratory syndrome (SARS) graphs, followed by using the Autoregressive Integrated Moving Average model, Autoregressive Moving Average model, Seasonal Autoregressive Integrated Moving-Average with Exogenous Regressors, and Holt Winter Rsquo;s Exponential Smoothing to predict infections, deaths, and GDP in China[30].

### Social Changes

17 papers were found having ***Social distancing*** keyword in their title summary of abstract of these papers are:

- Effectively implementing social distancing may be a challenge in informal settlements due to their density[33].
- Some social and behavioral research activities (e.g. data analysis, manuscript preparation) can be continued from other environments with appropriate security protocols in place[32].
- dwelling outlines for informal settlements in the city of Cape Town to demonstrate that with a 2 m measure, effective social distancing will be challenging.,The novel coronavirus has upended many traditional research procedures as universities and other research entities have closed to activate social distancing[33].

### Education

22 papers were found having ***Education*** keyword in their title summary of abstract of these papers are :

- several innovative solutions including the flipped classroom model, online practice questions, teleconferencing in place of in-person lectures, involving residents in telemedicine clinics, procedural simulation, and the facilitated use of surgical videos[34].
- Although there is no substitute for hands-on learning through operative experience and direct patient care, these may be ways to mitigate the loss of learning exposure during this time[34]
- In light of the recent coronavirus pandemic, there has never been more urgency to establish opportunities for supplemental online learning[35].

## Discussion

Going forward, COVID-19 is expected to have a significant impact on health systems, society and policy. Most of these sectors are currently scrambling to make better decisions in the light of evidence. While many countries have rolled out their plans over the next 12-18 months, the nature of the pandemic requires dynamic yet evidence-based decisions. Trends in peer-reviewed literature offer some degree of equipoise that is greater than daily news, hence knowledge synthesis from this line may prevent knee-jerk reactions in treatment and policy. A striking example of this dilemma has been the recommendation for Hydroxychloroquine that has gone from an optimistic to a negative narrative[46]. A similar controversy is currently emerging for the use of Remdesivir which sparked by the brief flashing of negative results on the WHO website which were then taken off. Such incidences indicate the extraordinary pressure to find evidence in the current circumstances. Our study is a small effort in plugging this gap by allowing human-in-the loop interaction with the published literature through the use of natural language processing, machine learning and visualization.

### Strength, limitations and future work

This study provides a model and an interface for the clinicians, researchers and policymakers to extract relevant information in the face of confusion that surrounds COVID-19. We used only peer reviewed articles in our analysis to make sure that information is validated to some extent, also realizing that even the peer-reviewed literature is currently conflicted. Another limitation of the study is the availability of only the abstracts on the WHO resource and the relatively small size of the resource. Future work in this direction will include full-texts of the available peer reviewed articles, primarily for the purpose of better model tuning. We will also create sections on our dashboard for knowledge synthesized by including pre-prints, expert summaries of social media posts of expert organizations and people, Government & Ministry Reports, WHO Reports and various other resources for improved inference in terms of data resources.

## Data Availability

All the data used in this study are publicly available from the WHO Covid-19 Global Literature on coronavirus disease maintained at https://search.bvsalud.org/global-literature-on-novel-coronavirus-2019-ncov/. Our analysis and the interactive resource CovidNLP is publicly available in a user friendly fashion at http://covidnlp.tavlab.iiitd.edu.in

https://search.bvsalud.org/global-literature-on-novel-coronavirus-2019-ncov/

http://covidnlp.tavlab.iiitd.edu.in

## Acknowledgements

This work was partially supported by the Wellcome Trust/DBT India Alliance Fellowship IA/CPHE/14/1/501504 awarded to Tavpritesh Sethi and the Center for Artificial Intelligence at IIIT-Delhi. Tavpritesh Sethi and Pradeep Singh also acknowledge the support received from the Department of Science and Technology vide project DST/INT/ISR/P-21/2017. Authors also acknowledge Prof. Rakesh Lodha from All India Institute of Medical Sciences, New Delhi, for his valuable inputs.

